# Antimicrobial resistance in WHO priority bacteria from a One Health perspective in Cameroon: a systematic review and meta-analysis

**DOI:** 10.64898/2026.04.03.26350076

**Authors:** Patrice Landry Koudoum, Winny Dora Ateudjieu, Adrien Nana, Giresse Wilfried Guemkam, Gauis Nditemeloung, Jerry Vladimir Abena, Esoh Rene Tanwieh, Njeodo Njongang Vigny, Teme Joseph Magloire, Aurelia Djeumako Mbossi, Joseph Kamgno, Hortense Gonsu Kamga

## Abstract

Antimicrobial resistance (AMR) is classified by the World Health Organization (WHO) as one of humanity’s ten global public health threats. This review aimed to estimate the prevalence, temporal trends and regional distribution of AMR in WHO priority bacteria across human, animal and environmental sources in Cameroon. This review was conducted following PRISMA 2020 guidelines, with the protocol registered in PROSPERO. A systematic literature search was conducted in Google Scholar, PubMed, African Journals Online, Hinari, and Africa indexus Medicus. Random effects models were used to estimate pooled prevalence and 95% confidence intervals (CIs), with subgroup analyses by bacterial source, region, and sampling period. Of 1566 articles screened, 115 met the inclusion criteria. The reported data encompassed 16 bacteria-antibiotic combinations in 16,948 isolates. Globally, third-generation cephalosporin (3GC) resistance in *E. coli* was the most prevalent (49.0%, 95% CI: 39.0-60.0%, I^2^=97.7%), reaching 77.0% (95% CI: 46.0-98.0%, I^2^=95.6%) in environmental isolates. The pooled prevalence of ESBL production in all included *Enterobacterales* was 37.0% (95% CI: 30.0-45.0%). Most of the highest resistance rates were observed in the Littoral region. The resistance rates between 2016 and 2025 were significantly higher than those from 2000 to 2015. These increases were more marked in fluoroquinolone-resistant *Salmonella* spp (1.0% to 48.0%, I^2^=97.3%, p<0.001), carbapenem-resistant *E. coli* (0% to 15%, I^2^=93.5%, p<0.001), and 3GC-resistant *E. coli* (34.0% to 64.0%, I^2^=97.6%, p=0.003). Antimicrobial resistance in WHO priority bacteria in Cameroon is high, unevenly distributed across regions and significantly increasing over time. These results underscore the crucial need for strengthened AMR surveillance to curb the growing threat of AMR in Cameroon.

## 1. Introduction

Antimicrobial resistance (AMR) is currently classified by the World Health Organization (WHO) as one of humanity’s ten global public health threats [1]. In 2019, antibiotic-resistant infections were responsible for 4.95 million deaths (95% UI: 3.62-6.57 million) [2]. Of these, 1.27 million deaths (0.911-1.71) were directly attributable to bacterial AMR [2]. This death toll is higher than that of malaria (643,000) [3] or AIDS (864,000) [4] during the same year. However, despite its high burden, AMR seems not to have received commensurate attention. The burden of AMR is disproportionately distributed worldwide, with resource-limited settings bearing the greatest burden. According to the systematic analysis by Murray et al (2021), the highest AMR burden was in sub-Saharan Africa, with an estimated mortality rate of 98.9 per 100,000 in 2019 [2].

Several factors promote the emergence and spread of AMR in resource-limited settings, including inadequate access to water, sanitation and hygiene, sub-optimal infection prevention and control, the high availability of antibiotics over-the-counter, the misuse of antibiotics in human and veterinary medicine and inadequate antimicrobial resistance detection in laboratories [5]. As the drivers of AMR are multisectoral and multifaceted, the One Health approach has been proposed as a holistic strategy to tackle AMR, as it acknowledges the interconnectedness of the human, animal and environmental interfaces. Antibiotic-resistant bacteria, resistance genes and mobile genetic elements carrying resistance genes can be spread within and across the three One Health compartments. As such, sustainable measures to address AMR should not only consider humans but the One Health triad [6].

Antimicrobial resistance surveillance systems are important tools for AMR containment as they help generate critical evidence for policy and decision-making in infection control and antibiotic usage guidelines in human and veterinary medicine [7]. Moreover, robust AMR surveillance systems can provide valuable data on the emergence, spread and prevalence of AMR. Therefore, understanding the local epidemiology of AMR is crucial to inform tailored strategies that effectively curb the growing AMR rates. Unfortunately, routine AMR surveillance is largely insufficient in most low- and middle-income countries, and the capacity for AMR monitoring remains inadequate in a significant number of African countries [8]. Cameroon adopted a National Action Plan (NAP) for the control of AMR in May 2018 for implementation between 2018 and 2020. However, no significant progress was made for its implementation and in March 2024, a revised action plan was published for implementation between 2024 and 2028. Much effort is still needed for standardized antimicrobial susceptibility testing, surveillance and prevention in the country [8, 9].

Among resistant bacteria causing deadly infections in humans, the WHO has listed carbapenem-resistant and third-generation cephalosporin-resistant *Enterobacterales,* such as *K. pneumoniae* and *E. coli,* as critical bacteria for the surveillance and development of new antibiotics due to their high disease burden [10]. A meta-analysis by Matakone and colleagues reported up to 50% resistance to third-generation cephalosporins in *E. coli* and *Klebsiella* spp, causing bloodstream infections in Cameroon [11]. Similarly, some studies have shown high rates of AMR in humans [12–18] and animals [19, 20] in Cameroon. A previous meta-analysis by Mouiche and collaborators in 2019 reported pooled AMR prevalence in the humans-animals-environment interface [21]. However, due to the increasing AMR trend, it is crucial to provide updated AMR prevalence to tailor specific interventions to help curb AMR in the country. Thus, this review aimed to estimate the prevalence, temporal trends and regional distribution of antimicrobial resistance in WHO priority bacteria across human, animal and environmental sources in Cameroon.

## 2. Methods

This study was designed as a systematic review with meta-analysis following the Preferred Reporting Items for Systematic Reviews and Meta-Analysis (PRISMA) 2020 guidelines [22]. The protocol was registered in PROSPERO (**CRD420251004416**) before the implementation of the study [23].

### 2.1. Search strategy and study selection

Two researchers independently conducted the literature search. A systematic search was carried out in the following electronic databases: Google Scholar, MEDLINE (via PubMed), African Journals Online (AJOL), Hinari, and African Index Medicus. Reference lists of relevant articles were also searched for additional articles. The publication date spanned from January 1, 2000, to August 7, 2025. Boolean operators “OR” and “AND” were used to identify articles on antibiotic resistance in relevant WHO priority bacteria [10], isolated from humans, animals, and the environment in Cameroon. *V. cholerae* was also included in the medium priority group, although it does not appear on the 2024 WHO priority list, due to recurrent cholera outbreaks in Cameroon [24]. The keywords used are presented in **Supplementary Figure 1.**

We included all original studies with abstracts and full texts available in English or French; studies using sound methodology for the identification of bacteria (culture, biochemical, serological and molecular methods) and studies that used a sound methodology for Antimicrobial Susceptibility Testing (AST) of bacteria in compliance with international standards, including: MIC, E-test, disk-diffusion methods. We excluded studies presenting aggregate data on antimicrobial resistance, such as those on “Enterobacteriaceae”, “Gram-positive” and “Gram-negative bacteria”; those reporting aggregate data on AMR in bacteria isolated at two or all three one health interfaces; and those reporting data on the aggregated family of antibiotics.

### 2.2. Citation management

All the resulting citations from the literature research were imported to the Rayyan platform for screening [25]. PLK, WDA and ANK independently excluded duplicates, screened the titles and abstracts for initial eligibility, and read the full texts of the articles for final inclusion. Resulting citations were then exported to EndNote software (Version 20). Any conflicts regarding the inclusion of an article were resolved by a group consensus.

### 2.3. Quality appraisal

To mitigate all potential threats to internal validity, each included study was assessed independently by two reviewers (PLK and WDA) using the Joanna Briggs Institute (JBI) checklist for cross-sectional studies [26]. For each criterion, each study was attributed a point for a “yes”. A final quality score was computed by summing the total number of points and fell between zero and eight. Studies attaining a score of ≥75% were designated as high quality, those scoring 50–74% were categorized as moderate quality, whilst studies falling below 50% were classified as low quality.

### 2.4. Data extraction

Data were extracted independently using Microsoft Excel 2016 by two reviewers (PLK and WDA), and any arising conflicts were resolved by group consensus. Extracted data included first author, year of publication, region of study, duration of study, study design, and sample size. Additional metadata collected included: patient type (inpatients/outpatients), age of patients, sample types, animal species, sample type and sampling site. We also extracted data on the methods used for bacteria identification, AST guidelines used for resistance breakpoints, the proportion of antibiotic - resistant bacteria, and the number of multidrug-resistant bacteria defined as resistance to at least three antimicrobial classes [21].

### 2.5. Data analysis

Descriptive statistics were performed using SPSS software (version 27), and meta-analysis was performed using Stata (version 15). QGIS software was used to generate maps. Publication bias was assessed using Egger’s regression [27] and Begg’s rank correlation [28] tests. A p-value less than 5% indicated statistically significant evidence for publication bias. The funnel plot was used to visualize publication bias. In case of publication bias, the fill and trim method was used to account for it [29]. The Cochrane Q statistic (significance level at p-value < 0.100) was used to evaluate heterogeneity between the studies, while the I^2^ statistic was used to quantify the heterogeneity [30]. I^2^ levels of 25%, 50%, and 75% indicated low, medium and high heterogeneity respectively [31].

Due to the high heterogeneity between articles, random effects models (REM) were used to determine the pooled prevalence of AMR per bacterium-antibiotic. The pooled prevalence was calculated if at least three studies reported a particular bacterium-antibiotic combination. The Freeman-Tukey double arcsine transformation was applied to the data due to the observed high variance between studies [32]. The total number of isolates tested for bacterium-antibiotic combination and the number of resistant isolates were used to generate effect sizes and standard errors, along with their 95% confidence intervals. Effect sizes and standard errors were used to determine pooled prevalence with their 95% confidence intervals. Subgroup meta-analysis was performed by bacterial source (human, animal, or environment), sampling year, and region.

## 3. Results

## 3.1. Search results

Systematic searching was conducted in five selected electronic databases and yielded a total of 1566 articles from PubMed, Google Scholar, Hinari, African Journals Online and Africa indexus Medicus. Twenty-two articles were identified through manual research. After the removal of duplicates and non-relevant articles, 141 articles were assessed for eligibility and 115 were finally included **(Figure 1).**

**Figure 1:**
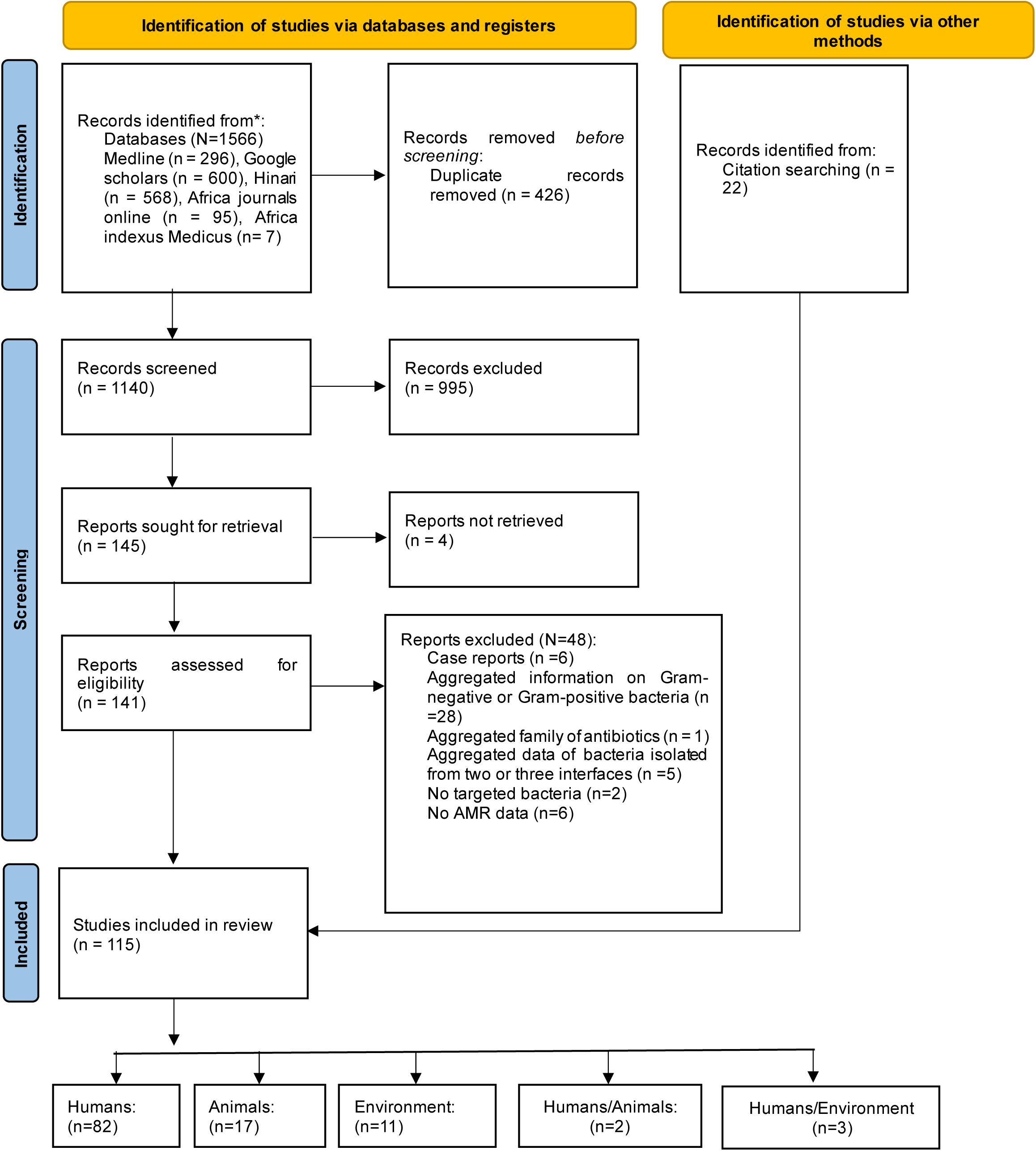
PRISMA flowchart of identified, screened and included studies.

### 3.2. Study characteristics

More than half (60.9%, 70/115) of the included studies were published between 2020 and 2025 **(Supplementary Figure 2).** Of the 115 studies included (**Figure 1**), the majority were cross-sectional studies (98.3%, 113/115), with samples collected prospectively in 84.3% (97/115). The studies included contained data from the 10 regions of Cameroon, with the largest proportions conducted in the Centre (38.3%, 44/115) and Littoral (20.0%, 23/115) regions (**Figure 2**).

**Figure 2:**
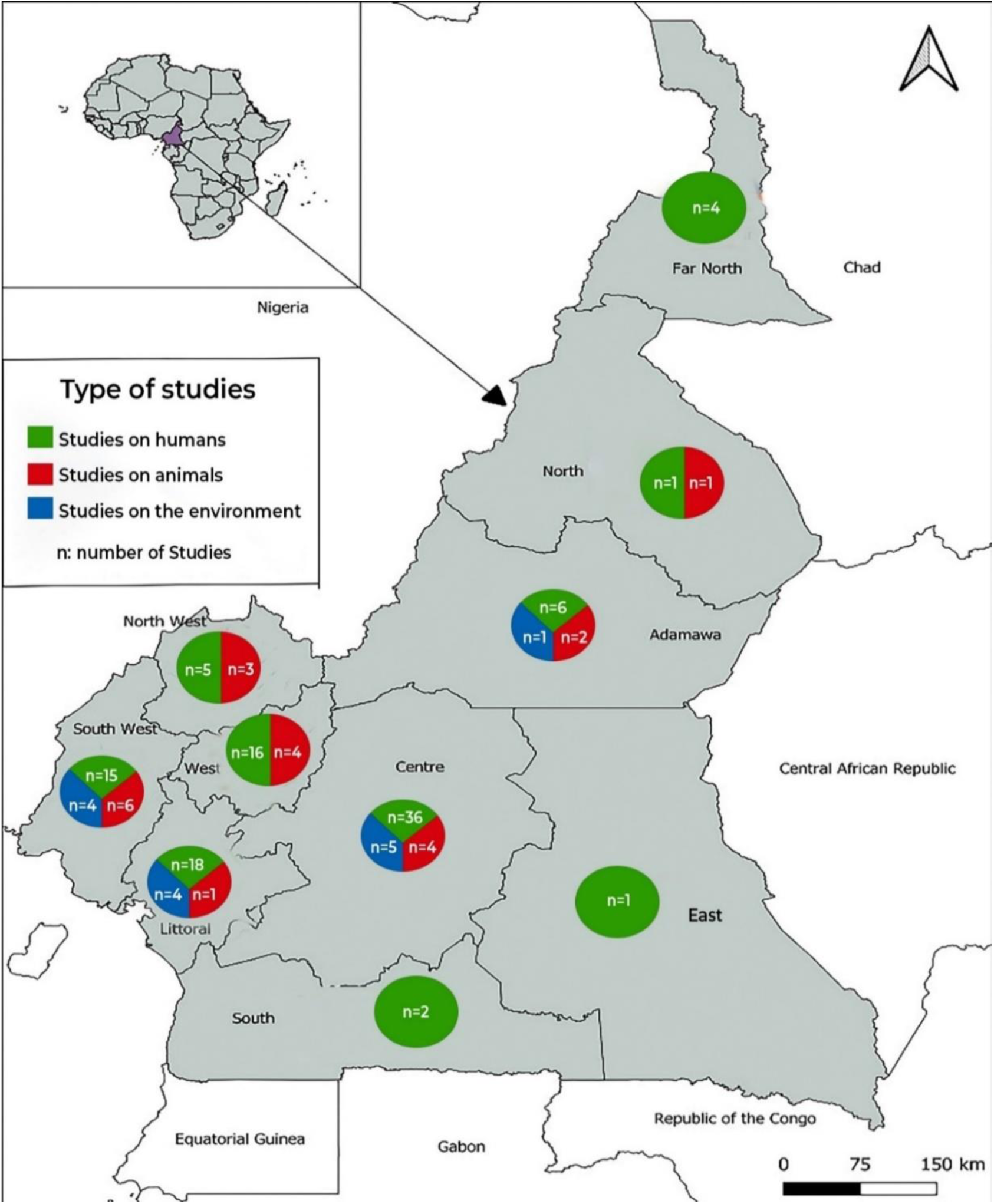
Map of Cameroon showing number of articles included per region.

Quality assessment showed that a majority of articles were of low risk (82.6%, 95/115) (**Table I**).

**Table I:** Distribution and characteristics of studies included in the review.

| Characteristics | Overall<br>(n=115) | Human<br>studies<br>(n=87) | Animal<br>studies (19) | Environment<br>studies (n=14) |
| --- | --- | --- | --- | --- |
| **Language** |  |  |  |  |
| French | 3 (2.61) | 3 (3.44) | 0 | 0 |
| English | 112 (97.39) | 84 (96.55) | 19 (100) | 14 (100) |
| **Study design** |  |  |  |  |
| Longitudinal | 2 (1.74) | 2 (2.29) | 0 | 0 |
| Cross sectional | 113 (98.26) | 85 (97.70) | 19 (100) | 14 (100) |
| **Sample collection** |  |  |  |  |
| Prospective | 97 (84.35) | 71 (81.61) | 17 (89.47) | 14 (100) |
| Retrospective | 16 (13.91) | 14 (16.09) | 2 (10.53) | 0 |
| Prospective and retrospective | 2 (1.74) | 2 (2.29) |  |  |
| **Quality** |  |  |  |  |
| High | 95 (82.61) | 70 (80.46) | 17 (89.47) | 12 (13.79) |
| Moderate | 18 (15.65) | 15 (17.24) | 2 (10.53) | 2 (14.29) |
| Low | 2 (1.74) | 2 (2.29) |  |  |
| **Regions from which bacteria were isolated** |  |  |  |  |
| Adamawa | 9 (7.83) | 6 (6.89) | 2 (10.53) | 1 (7.14) |
| North | 2 (1.74) | 1 (1.15) | 1 (5.26) | 0 |
| Far-North | 4 (3.48) | 4 (4.60) | 0 | 0 |
| Centre | 44 (38.26) | 36 (41.38) | 4 (21.05) | 4 (28.57) |
| Littoral | 23 (20.00) | 18 (20.69) | 1 (5.26) | 4 (28.57) |
| West | 20 (17.39) | 16 (18.39) | 4 (21.05) | 0 |
| South | 2 (1.74) | 2 (2.30) | 0 | 0 |
| East | 1 (0.87) | 1 (1.15) | 0 | 0 |
| South-West | 24 (20.87) | 15 (17.24) | 6 (31.58) | 3 (21.43) |
| North-West | 8 (6.96) | 5 (5.75) | 3 (15.79) | 0 |
| **Human population only** |  |  |  |  |
| Healthy |  | 9 (10.34) |  |  |
| PLWHIV |  | 6 (6.89) |  |  |
| Patients with chronic disease |  | 5 (5.75) |  |  |
| Medical personnel |  | 2 (2.29) |  |  |
| Cancer patients |  | 4 (4.60) |  |  |
| Farmers |  | 2 (2.29) |  |  |
| Pregnant women |  | 4 (4.60) |  |  |
| Other patients |  | 55 (63.22) |  |  |
| **Animal population only** |  |  |  |  |
| Bovine |  |  | 8 (42.11) |  |
| Poultry |  |  | 7 (36.84) |  |
| Shrimp |  |  | 1 (5.26) |  |
| Swine |  |  | 2 (10.53) |  |
| Fish |  |  | 4 (21.05) |  |
| **Environmental population only** |  |  |  |  |
| Well water |  |  |  | 4 (28.57) |
| Banknotes and coins |  |  |  | 1 (7.14) |
| Poultry litter |  |  |  | 1 (7.14) |
| Wastewater |  |  |  | 2 (14.28) |
| Inanimate hospital surfaces and medical devices |  |  |  | 6 (42.86) |

PLWHIV= Patients Living with HIV

Eighty-two studies focused on only humans (71.3%, 82/115), 17 on only animals (14.8%, 17/115), 11 on only the environment (9.6%, 11/115), two (1.4%, 2/115) on both humans and animals and three (2.6%, 3/115) on both humans and the environment. Studies with the human population consisted mainly of patients with infections (82.8%, 72/87). The animal population consisted mainly of bovine (42.1%, 8/19) and fish (21.1%, 4/19), while in environmental studies, the main samples were hospital surfaces (42.9%, 6/14) and well water (28.6%, 4/14) (**Table I**).

#### 3.2.1. Microbiological characteristics of included studies

A total of 16 WHO priority pathogens (including *Vibrio cholerae*) were identified with the most frequent being *E. coli* (65.2%, 75/115), *S. aureus* (35.7%, 41/115), *S. enterica* (23.5%, 27/115), *K. pneumoniae* (21.7%, 25/115) and *P. aeruginosa* (16.5%, 9/115) (**Supplementary Figure 3**).

The most commonly used technique for bacterial species identification was Analytical Profile Index (API) galleries (59.1%, 68/115), followed by biochemical tests (20.9%, 24/115) and PCR (6.1%, 7/115).

Regarding AST, disk diffusion (90.4%, 104/115) and MIC determination (12.2%, 14/115) were the most used methods. Altogether, 58 (50.4%) studies performed quality control of AST, with 57 (49.6%) studies using American Type Culture Collection (ATCC) reference strains (**Supplementary Table I**). The interpretation of AST results was based on European Committee on Antimicrobial Susceptibility Testing (EUCAST) guidelines (51.3%, 59/115) and Clinical and Laboratory Standards Institute (CLSI) guidelines (43.5%, 50/115). Resistance gene detection was reported in 34 studies.

### 3.3. Pooled prevalence of antibiotic resistance in the human, animal and environmental interfaces

This meta-analysis reports 16 bacteria-antibiotic combinations in 16,948 WHO priority bacterial isolates from Cameroon between 2000 and 2025, with the most commonly reported being 3GC-resistant *E. coli* (n=4392) and Methicillin-Resistant *S. aureus* (MRSA) (n=3,703) (**Supplementary Figure 4**).

#### 3.3.1. Pooled prevalence of antibiotic resistance in critical priority bacterial pathogens

The pooled prevalence of 3GC resistance in *E. coli* was 49.0% (95% CI: 39.0-60.0%, I^2^=97.7%) (**Figure 3**), with the highest prevalence observed in the environmental interface (77.0%, 95% CI: 46.0-98.0%, I^2^=95.6%), although there was no significant difference across the three One Health interfaces (**Figure 4**).

**Figure 3:**
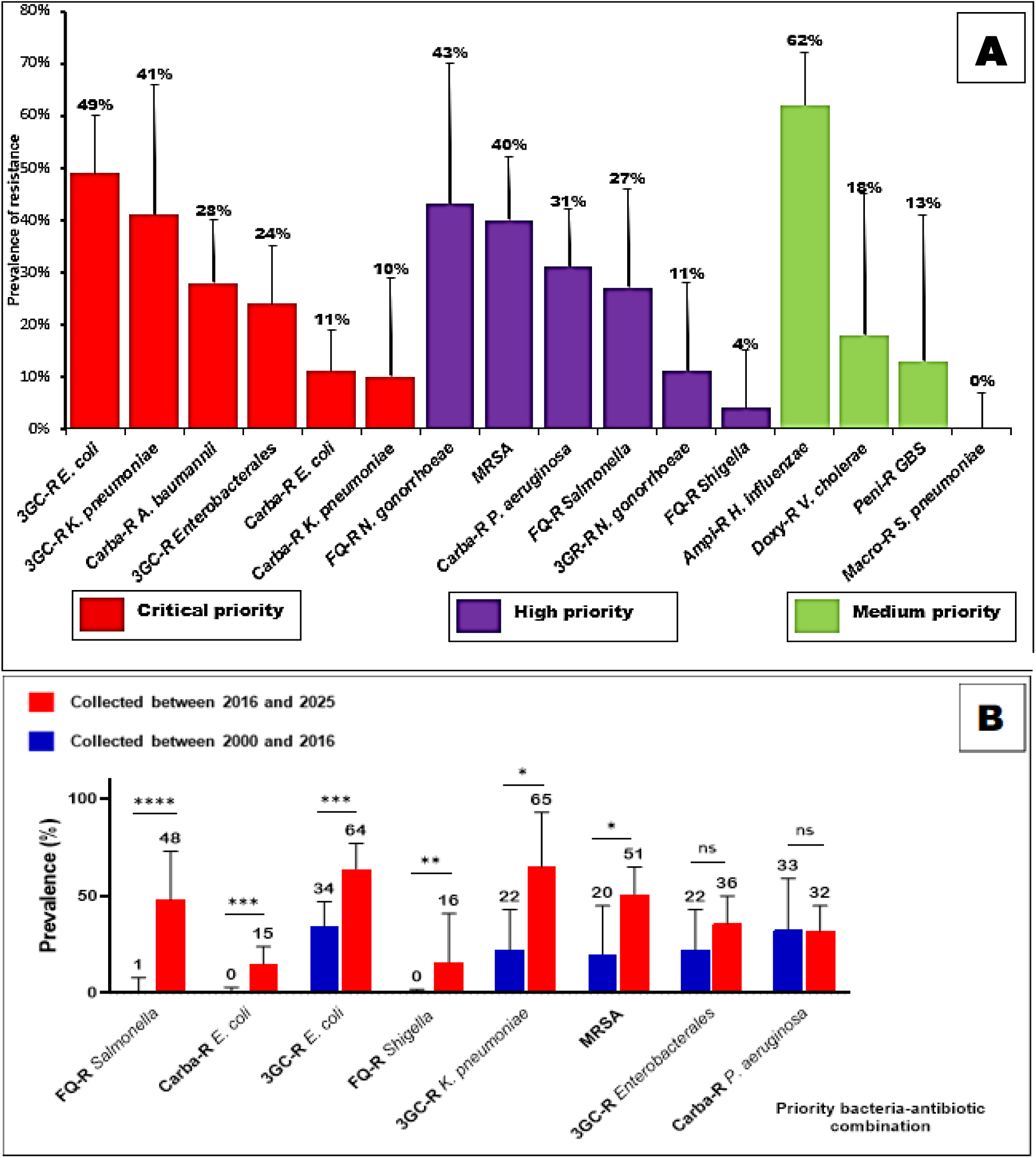
Pooled prevalence of antibiotic resistance in **(A)** WHO priority bacterial pathogens across One Health interfaces and **(B)** sampling year. 3GC-R=Third-generation Cephalosporin Resistant, MRSA=Methicillin Resistant *S. aureus*, Carba-R=Carbapenem-Resistant, FQ-R=Fluoroquinolone-Resistant, Ampi-R=Ampicillin-Resistant, Doxy-R=Doxycycline-Resistant, Macro-R=Macrolide-Resistant, Peni-R=Penicillin-Resistant. **** =p<0000.1, ***=p<0.0001, **=p<0.001, *=p<0.01, ns=not significant.

**Figure 4:**
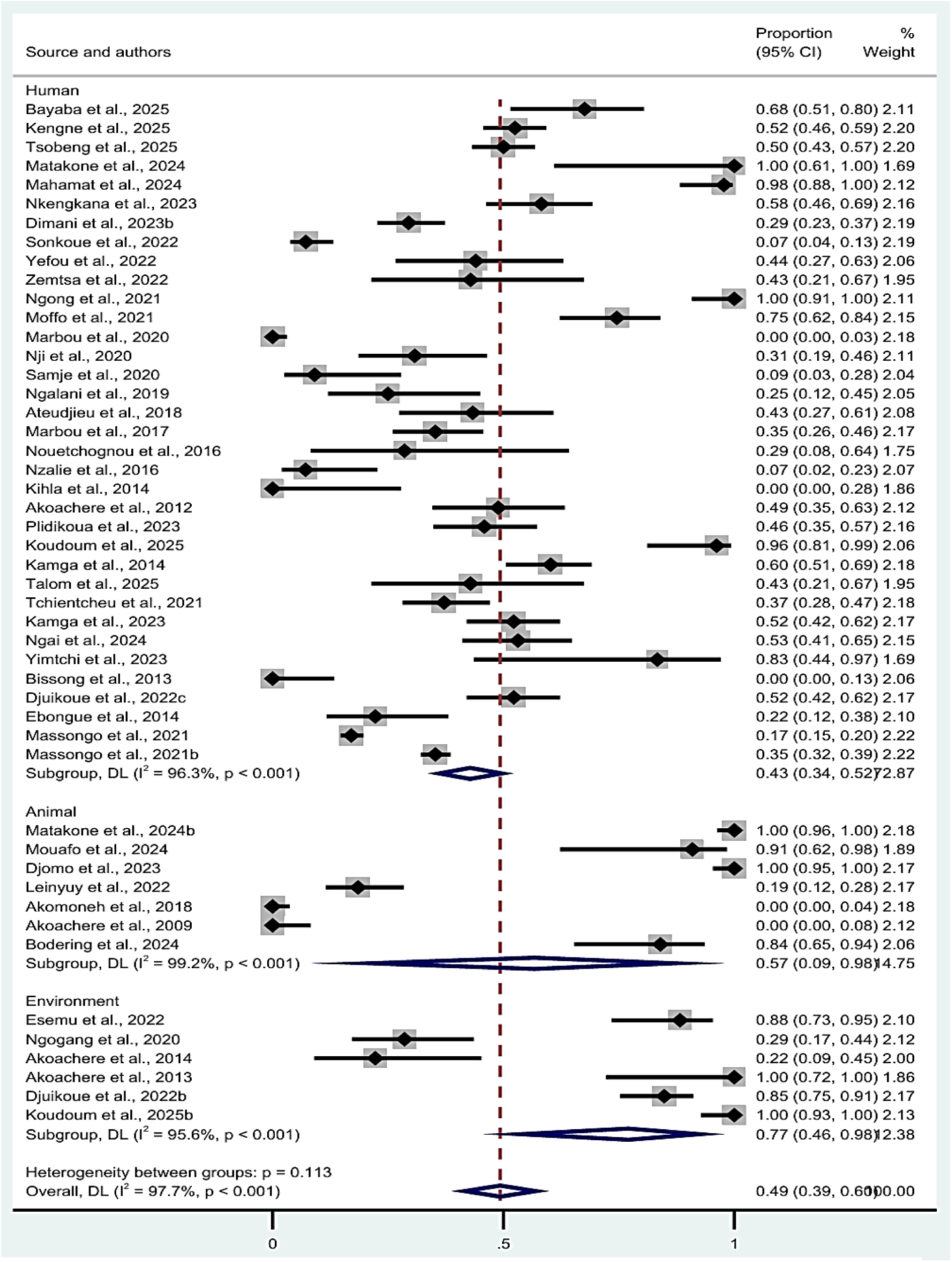
Forest plot of pooled prevalence of third-generation cephalosporin resistance in *E. coli* with subgroup meta-analysis by source. Note: weights and between subgroup analysis are from random effect models.

Similarly, the overall pooled prevalence of carbapenem resistance in *E. coli* was 11.0% (95% CI: 5.0-19.0%, I^2^=93.7%), which was again highest in the environmental interface (40.0%, 95% CI: 0-97.0%, 97.9%), with no significant difference across the three interfaces (**Supplementary Figure 5**). The overall pooled prevalence of 3GC resistance in *K. pneumoniae* across three interfaces was 41.0% (95% CI: 19.0-66.0%, I^2^=95.9%) (**Supplementary Figure 6A**) while the pooled prevalence of carbapenem resistance in *K. pneumoniae* was 10% (95% CI: 0-29.0% I^2^=94.3%) (**Supplementary Figure 6B**). Carbapenem-resistant *A. baumannii* was only reported by human studies with a pooled prevalence of 28.0% (95% CI: 17.0-40.0%, I^2^=88.7%). Regarding other *Enterobacterales* (*Citrobacter* spp., *Proteus* spp., *Serratia* spp., *Enterobacter* spp. and *Morganella* spp.), the overall pooled prevalence of 3GC resistance was 24% (95% CI: 14.0-35.0%, I^2^=89.1%) (**Supplementary Figure 7**). The pooled prevalence of ESBL production in all included *Enterobacterales* was 37.0% (95% CI: 30.0-45.0%).

#### 3.3.2. Pooled prevalence of antibiotic resistance in high-priority bacterial pathogens

The overall pooled prevalence of fluoroquinolone resistance in *Salmonella* spp. was 27% (11.0-46.0%, I^2^=97.1%) with no statistically significant difference across the three interfaces (**Supplementary Figure 8A**). Fluoroquinolone resistance in *Shigella* spp. across all interfaces was low (4.0%, 95% CI: 0-15.0, I^2^=88.8%). Carbapenem resistance in *P. aeruginosa* was mainly observed in human isolates with a pooled prevalence of 32% (95% CI: 21.0-42.0, I^2^=92.2%) (**Supplementary Figure 8B**). In humans, the pooled prevalence of fluoroquinolone and 3GC resistance in *N. gonorrhoeae* was respectively, 43% (95% CI: 18-70%, I^2^=97.9) and 11.0% (95% CI: 1.0-28.0%, I^2^=96.4%). The pooled prevalence of methicillin resistance in *S. aureus* isolates was 40.0% (95% CI: 29.0-52.0%, I^2^=97.7%), with the highest prevalence observed in humans, 46.0% (33.0-58.0%, I^2^=97.6%) (**Supplementary Figure 9**).

#### 3.3.3. Pooled prevalence of antibiotic resistance in medium priority bacterial pathogens

Macrolide resistance in *S. pneumoniae* was tested in two human studies that reported a prevalence of 0% (95% CI: 0-7.0%) and 30% (95% CI: 17.0-48.0). Penicillin resistance in GBS was reported in two human and one animal study and resulted in a pooled prevalence of 13% (95% CI: 0-41.0%, I^2^=82.9%). Ampicillin resistance in *H. influenzae* was reported in three studies with a pooled prevalence of 62% (95% CI:51.0-72.0, I^2^=52.2%). Doxycycline resistance in *V. cholerae* was reported by 3 environmental and one human study, resulting in a pooled prevalence of 18% (95% CI: 1.0-45.0, I^2^=92.2%).

### 3.4. Pooled prevalence of antibiotic resistance in priority bacterial pathogens according to the region of bacterial isolation

Significant disparities in the prevalence of antibiotic resistance according to the region of bacterial isolation were observed for 3GC resistance in *K. pneumoniae* (I^2^=95.9%, p<0.0001) (**Figure 5**), 3GC resistance in *E. coli* (I^2^=97.7%, p=0.001), 3GC-resistant *Enterobacterales* (I^2^=89.3%, p=0.007) (**Figure 5**), methicillin resistance in *S. aureus* (I^2^=96.6%, p=0.007) and Carbapenem resistance in *P. aeruginosa* (I^2^=92.8%, p=0.017) (**Supplementary Table II**). Globally, the highest resistance rates were observed in the Littoral and Centre regions, while the lowest were observed in the Southwest and Northwest regions. The highest resistance rates reported in the Littoral region included 3GC-resistant *Enterobacterales* (54.0%, 95% CI:30.0-77.0%), MRSA (62.0%, 95% CI:31.0-89.0%), and carbapenem-resistant *P. aeruginosa* (51.0%, 95% CI:26.0-75.0%). The Centre region showed the highest prevalence of 3GC-resistant *K. pneumoniae* (74.0%, 95% CI:38.0-98.0%), while the Adamawa region had the highest rates of 3GC-resistant (66.0%, 95% CI:24.0-98.0%) and carbapenem-resistant *E. coli* (55.0%, 95% CI:0-100.0%).

**Figure 5:**
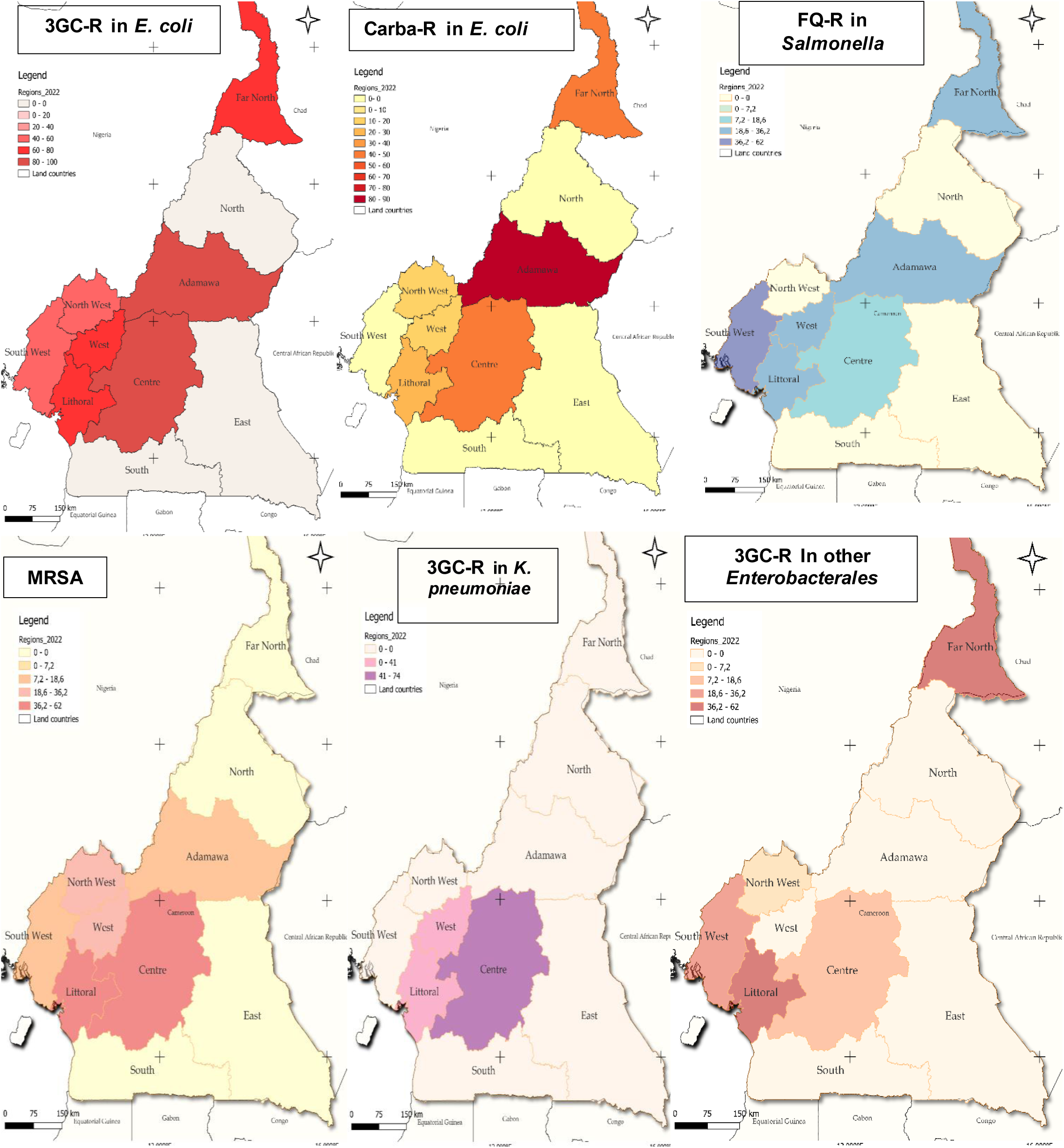
Regional variation in the pooled prevalence (in %) of antibiotic resistance.

### 3.5. Prevalence of antibiotic resistance in priority bacterial pathogens according to sampling period

For nearly all tested bacteria-antibiotic combinations (except for carbapenem-resistant *P. aeruginosa* and 3GC-resistant *Enterobacterales*), the resistance rates in isolates collected between 2016 and 2025 were significantly higher than those collected between 2000 and 2015 (**Supplementary Figure 10**). This difference was more marked in fluoroquinolone resistance in *Salmonella* spp. (1.0% vs 48.0%, I^2^=97.3%, p<0.00001) followed by carbapenem resistance in *E. coli* (0% vs 15%, I^2^=93.5%, p<0.00001) (**Figure 3**), 3GC resistance in *E. coli* (34.0% vs 64.0%, I^2^=97.6%, p=0.003), fluoroquinolone resistance in *Shigella* spp. (0% vs 16.0%, I^2^=93.5%, p=0.008), 3GC resistance in *K. pneumoniae* (22.0% vs 65.0%, I^2^=95.5%, p=0.030) and MRSA (20.0% vs 51.0%, I^2^=95.5%, p=0.036) (**Figure 3**).

## Discussion

Antimicrobial resistance (AMR) has emerged as a major threat to human health with profound clinical and economic implications [33]. While AMR prevalence data are crucial for developing containment strategies, it is still underreported for most sub-Saharan African countries (Cameroon included) [8]. To the best of our knowledge, this review represents the largest, most recent, and most comprehensive dataset on AMR in Cameroon across the One Health interfaces. Previous reviews did not include most WHO priority bacteria and presented aggregated AMR at the genus level [21], focused on a single infection in humans [34] or on a single resistance phenotype across One Health interfaces [35].

We synthesized data from 115 published articles reporting AMR in WHO priority bacterial pathogens isolated from humans, animals and the environment in Cameroon. Collectively, these studies reported 16 bacteria-antibiotic combinations in 16,948 priority bacterial isolates. Resistance data for *E. coli* was the most reported (n=4,392) followed by *S. aureus* (n=3,703) and *P. aeruginosa* (n=1,246). These pathogens are among the leading causes of AMR-related deaths in Africa [2]. In 2019, in Africa, antibiotic-resistant *E. coli* was associated with 147,000 (112,000–189,000) deaths, while antibiotic-resistant *S. aureus* was associated with 136,000 (109,000–172,000) deaths, and antibiotic-resistant *P. aeruginosa* was associated with 56,000 (42,000–72,000) deaths [2].

Our results indicate high AMR prevalence in WHO critical-, high- and medium-priority bacterial pathogens with a substantial variability across regions, time and One Health interfaces. Third-generation cephalosporin-resistant *E. coli* was the most reported pathogen-drug combination, with a pooled prevalence of 43.0% (95% CI: 34%-52%) in humans, 57% (95% CI: 9%-98%) and 77% (95% CI: 46%-98%) in the environment. The prevalence in human isolates exceeds estimates reported in most African countries, typically below 40% [2]. This elevated pooled prevalence is likely driven by the misuse and overuse of antibiotics, especially β-lactams (penicillin and cephalosporins) and β-lactams combination (Amoxicillin + clavulanic acid) in hospital and community settings in Cameroon [36]. Ekambi et al. (2019) reported that up to 50% of antibiotics are sold in private pharmacies without a medical prescription [36]. This situation is further worsened by the fact that, antibiotics are readily available over-the-counter in Cameroon [36]. The pooled prevalence of 3GC-resistant *E. coli* isolates from animals and the environment in our study is higher than that reported by a recent Ethiopian meta-analysis, which reported a prevalence of 41% (95% CI: 0–97%) in animals and 14% (95% CI: 0–41%) in the environment [32]. The higher prevalence of 3GC-resistant *E. coli* in environmental samples suggests that the environment serves as an important reservoir for AMR, playing a key role in resistance dissemination in Cameroon. Antibiotic-resistant bacteria and genes from humans and animals may persist in the environment and subsequently spread to other One Health components. These results reinforce the incorporation of surveillance of antibiotic-resistant human pathogens in the environment [37].

Carbapenem resistance in *A. baumannii* and *P. aeruginosa* was reported with pooled prevalences of 28% (95% CI: 17-40%) and 31% (95% CI: 23-45%), respectively in human isolates. These prevalences are higher than those reported by a recent meta-analysis of sub-Saharan African studies which found a prevalence of 20% (4-43%) and 8% (95% CI: 2-17%) for carbapenem resistance in *A. baumannii* and *P. aeruginosa* respectively. This is particularly worrying as carbapenems are considered last resort antibiotics in the case of multidrug-resistance [38]. Similarly, the overall pooled prevalence of MRSA was 40% (95% CI: 29-52%), exceeding rates reported in most African countries [2] and the 1.9% to 34.6% range documented by a recent One Health mapping review in Cameroon [35].

Subgroup analysis of resistance according to geographical region revealed significant disparities for five bacteria-antibiotic combinations: 3GC-resistant *E. coli*, Carbapenem resistance in *P. aeruginosa*, 3GC resistance in *K. pneumoniae* and 3GC resistance in *Enterobacterales* (including *Citrobacter* spp., *Proteus* spp., *Serratia* spp., *Enterobacter* spp. and *Morganella* spp.). Pooled resistance rates were higher for most bacteria-antibiotic combinations in the Littoral and Centre regions but lower in the Northwest and Southwest regions. This is likely because the largest cities of Cameroon are found in the Centre (Yaounde) and Littoral (Douala) regions. These metropolitan cities are characterized by higher accessibility to antibiotics both in the legal sector and over the counter. Thus, antibiotics can be readily purchased and consumed, triggering the emergence and selection of resistant pathogens [39]. In addition, most of the highest-rated hospitals are located in these two cities. [40]. Consequently, there is a higher number of critically ill patients with multidrug-resistant bacteria likely sampled in these regions [40]. The observed high prevalence may also reflect the overrepresentation of these two regions, as more than half of the included studies originated from these two regions.

Regarding antibiotic resistance according to bacterial sampling years, a significant increase in resistance between 2000-2015 to 2016-2025 was observed for six bacteria-antibiotic combinations with the most significant increase observed for fluoroquinolone resistance in *Salmonella* spp. (1.0% vs 48.0%, p<0.00001) followed by carbapenem resistance in *E. coli* (0% vs 15% p<0.00001) and 3GC resistance in *E. coli* (34.0% vs 64.0%, I^2^=97.6%, p=0.003). The rise in fluoroquinolone resistance in *Salmonella* spp. is worrying as fluoroquinolones are used as frontline treatments for typhoid fever. The highest pooled fluoroquinolone prevalence was found in animal isolates. This suggests the emergence and spread of fluoroquinolone resistance genes in animals which was most noticeable in poultry. This is likely due to the increase in the use of antibiotics in food animal production. Mouiche et al. reported that 217.67 tons of antimicrobials for veterinary use were imported between 2014 and 2019, with a 104% increase in import volume [41]. This is worrying as 34% of these antibiotics were critically important to human medicine (WHO AwaRe classification). Moreover, even antibiotics exclusively used in veterinary medicine, such as the fluoroquinolone enrofloxacin, can select for resistance to clinically relevant fluoroquinolones such as ciprofloxacin and emerging resistant strains may spread directly to humans or indirectly through shared environments [42]. The rise of carbapenem resistance in *E. coli* is also worrying. This is likely due to the increasing resistance of *E. coli* to 3GC (also observed in this study) via diverse mechanisms such as the production of enzymes, including ESBLs and cephalosporinases (AmpC). This rise likely prompted greater reliance on carbapenems, which in turn escalated carbapenem resistance.

This review reports high AMR across humans, animals and the environment in Cameroon, highlighting the intermingling of AMR drivers across One Health interfaces. Notwithstanding, this review had some limitations: First, data on AMR across three One Health interfaces were not available for all regions. Studies on the environment were present in 4 regions and animal studies in 7 of the 10 regions. This limitation precluded subgroup analysis of some bacteria-antibiotic combinations. Second, the variation in AST methods and interpretation guidelines likely reduced the comparability of results. Thirdly, nearly all the studies were conducted in urban areas, making it difficult to generalize the findings to rural settings. Lastly, several studies reported AMR data in bacterial isolates sampled over many years without reporting AMR data at each time point. This hindered their inclusion in subgroup meta-analysis by sampling period.

Despite these limitations, this study provides the most comprehensive synthesis to date of antimicrobial resistance in WHO priority bacteria in Cameroon. It covers a period of 25 years (2000–2025) and integrates a One Health approach, allowing a broader understanding of AMR dynamics. In addition, regional subgroup analyses highlight important geographic disparities in AMR distribution, identifying key hotspots such as the Littoral, Centre and Adamawa regions. These findings provide valuable evidence to inform targeted AMR surveillance strategies, antimicrobial stewardship programs and region-specific containment policies in Cameroon.

## Conclusion

Antimicrobial resistance in WHO priority bacteria in Cameroon is high and unevenly distributed across regions, with the Littoral region emerging as a major hotspot for 3GC-resistant *Enterobacterales*, methicillin-resistant *Staphylococcus aureus* (MRSA) and carbapenem-resistant *Pseudomonas aeruginosa*. Moreover, resistance rates have significantly increased over time, with the most notable increases reported for fluoroquinolone-resistant *Salmonella*, carbapenem-resistant *E. coli*, and 3GC-resistant *E. coli*. AMR rates were significantly higher in the environment for carbapenem-resistant *E. coli* and 3GC-resistant *Enterobacterales,* while MRSA was significantly higher in humans. These findings underscore the urgent need for an integrated One Health approach that combines strengthened surveillance and addresses environmental reservoirs to curb the rising threat of AMR in high-burden regions of Cameroon.

### CRediT authorship contribution statement

**Patrice Landry Koudoum**: Conceptualization, Data curation, Formal analysis, Methodology, Software, Writing – original draft, Writing – review and editing. **Winny Dora Ateudjieu**: Conceptualization, Data curation, Formal analysis Methodology, Software, Writing – review and editing. **Adrien Nana**: Conceptualization, Data curation, Formal analysis Methodology. **Giresse Wilfried Guemkam**: Methodology, Data curation, Software. **Gauis Nditemeloung**: Methodology, Data curation, Software. **Jerry Vladimir Abena**: Methodology, Data curation, Software, **Esoh Rene Tanwieh**: Methodology. **Njeodo Njongang Vigny**: Methodology. **Teme Joseph Magloire**: Software, Visualization. **Aurelia Djeumako Mbossi**: Methodology. **Joseph Kamgno**: Supervision, Writing – review and editing. **Hortense Gonsu**: Conceptualization, Supervision, Writing – review and editing.

## Funding

The authors received no specific funding for this study.

### Declaration of competing interest

The authors declare no conflict of interest.

## Data Availability

All data produced in the present study are available upon reasonable request to the authors

## Acknowledgments

The authors are grateful for the assistance in data analysis provided by Matakone Moise and Visualization by Ulrich Loic Ngopdop

## Notes

### Competing Interest Statement

The authors have declared no competing interest.

### Clinical Protocols

https://www.crd.york.ac.uk/PROSPERO/view/CRD420251004416

### Funding Statement

This study did not receive any funding

