## Supplementary Tables and Figures for "Antimicrobial resistance in WHO priority bacteria from a One Health perspective in Cameroon: a systematic review and meta-analysis"

**Supplementary Table I:** Characteristics of included studies on bacterial identification, antimicrobial susceptibility testing and resistance gene detection

| Characteristic | Number of studies n (%) N=115 |
|---|---|
| **Antimicrobial Susceptibility Testing technique** | |
| Disk diffusion | 104 (90.43) |
| Minimal Inhibitory Concentration (MIC) by broth dilution | 14 (12.17) |
| MIC by E-test | 2 (1.74) |
| ATB EU Gallery | 1 (0.87) |
| **Guideline for AST interpretation** | |
| European Committee for Antimicrobial Susceptibility Testing (EUCAST) | 59 (51.30) |
| Clinical Laboratory Standards Institute (CLSI) | 50 (43.48) |
| Not specified | 9 (7.83) |
| **Quality control of antibiotics** | |
| American Type Culture Collection (ATCC) | 57 (49.57) |
| *Collection de l'Institut Pasteur* (CIP) | 1 (0.87) |
| Not done | 57 (49.57) |
| **Bacteria identification techniques** | |
| Analytical Profile Index (API) | 68 (59.13) |
| Polymerase chain reaction | 7 (6.09) |
| Chromogenic agar | 3 (2.61) |
| Seroagglutination | 7 (6.09) |
| VITEK system | 6 (5.22) |
| Biochemical tests | 24 (20.87) |
| Enterosystem | 3 (2.61) |
| MALDI-TOF | 4 (3.48) |
| **Detection of resistance genes** | |
| Polymerase chain reaction | 31 (26.96) |
| Sequencing | 3 (2.61) |
| Not done | 81 (70.43) |

MALDI-TOF= Matrix-Assisted Laser Desorption/Ionization–Time Of Flight

**Supplementary Table II**: Regional variation in the pooled prevalence of antibiotic resistance

| **Bacteria-antibiotic combination** | **3GC-R *E. coli*** | **Carba-R *E. coli*** | **MRSA** | **FQ-R *cter*** | **3GC-R *Enterobacterales*** | **3GC-R *K. pneumoniae*** | **Carba-R *P. aeruginosa*** |
|---|---|---|---|---|---|---|---|
| $I^2$ (p-value) | 97.7% (0.001) | 93.7% (0.549) | 96.6% (0.007) | 97.2% (0.163) | 89.3% (0.007) | 95.9% (<0.0001) | 92.8% (0.017) |
| Adamawa | **66.0 (24.0-98.0)** | **55.0 (0-100.0)** | 17.0 (3.0-37.0) | 30.0 (0-100.0) | NA | NA | NA |
| Centre | 64.0 (47.0-80.0) | 10.0 (2-21.0) | 45.0 (27.0-64.0) | 9.0 (2.0-19.0) | 15.0 (0-40.0) | **74.0 (38.0-98.0)** | **20.0 (12.0-28.0)** |
| Far-North | 43.0 (27.0-61.0) | 10.0 (3.0-26.0) | NA | 29.0 (8.0-64.0) | NA | NA | NA |
| Littoral | 54.0 (30.0-78.0) | **4.0 (1.0-9.0)** | **62.0 (31.0-89.0)** | 21.0 (0-88.0) | **54.0 (30.0-77.0)** | 34.0 (8.0-66.0) | **51.0 (26.0-75.0)** |
| North-West | **9.0 (3.0-28.0)** | 7.0 (3.0-16.0) | 34.0 (28.0-41.0) | NA | 50.0 (19-81.0) | NA | NA |
| South-West | 24.0 (0-65.0) | NA | **12.0 (0-49.0)** | **7.0 (0-27.0)** | **6.0 (0-21.0)** | **0 (0-3.0)** | NA |
| West | 41.0 (20.0-64.0) | 8.0% (0-31.0) | 21.00 (14.0-29.0) | **46.0 (15.0-78.0)** | 31.0 (18.0-46.0) | 41.0 (24.0-58.0) | NA |

3GC-R=Third-generation Cephalosporin Resistance, MRSA=Methicillin Resistant *S. aureus*, Carba-R=Carbapenem-Resistant, FQ-R=Fluoroquinolone-Resistant. Colored in green: lowest prevalence, Colored in red: highest prevalence.

(Antimicrobial resistance* OR AMR OR antibiotic resistance OR multiresistant OR multi-drug resistant OR MDR OR drug resistance OR bacterial resistance) **AND** (Acinetobacter baumannii OR Escherichia coli OR E. coli OR Klebsiella pneumoniae OR Salmonella OR Pseudomonas aeruginosa OR Shigella OR Enterobacter OR Serratia OR Proteus OR Citrobacter OR Morganella OR Enterococcus OR Neisseria OR Staphylococcus aureus OR S. aureus OR Methicillin-resistant OR MRSA OR S. pneumoniae OR Haemophilus influenza OR Group B streptococcus OR S. agalactiae OR GBS OR S. pyogenes OR Group A streptococcus OR GAS OR Vibrio cholerae OR Neisseria gonorrhoeae) **AND** (Cameroon OR Yaoundé OR Douala OR Bafoussam OR Adamawa OR Ngaoundere OR Garoua OR Bertoua OR Ebolowa OR Maroua OR Buea OR Bamenda)

**Supplementary Figure 1:** Keywords used for literature search in PubMed.

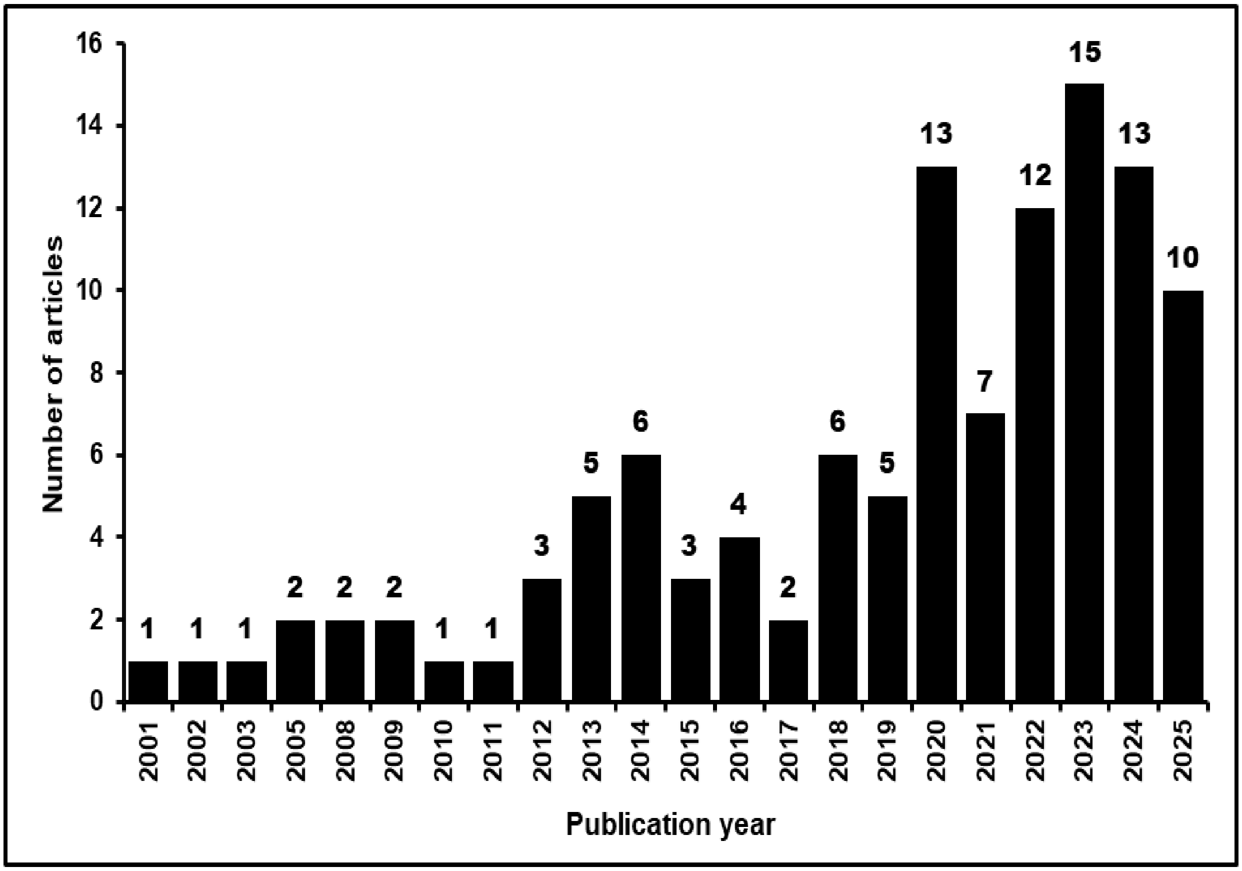


**Supplementary figure 2:** Number of studies reporting antibiotic resistance in Cameroon published per year from 2000 to 2025.

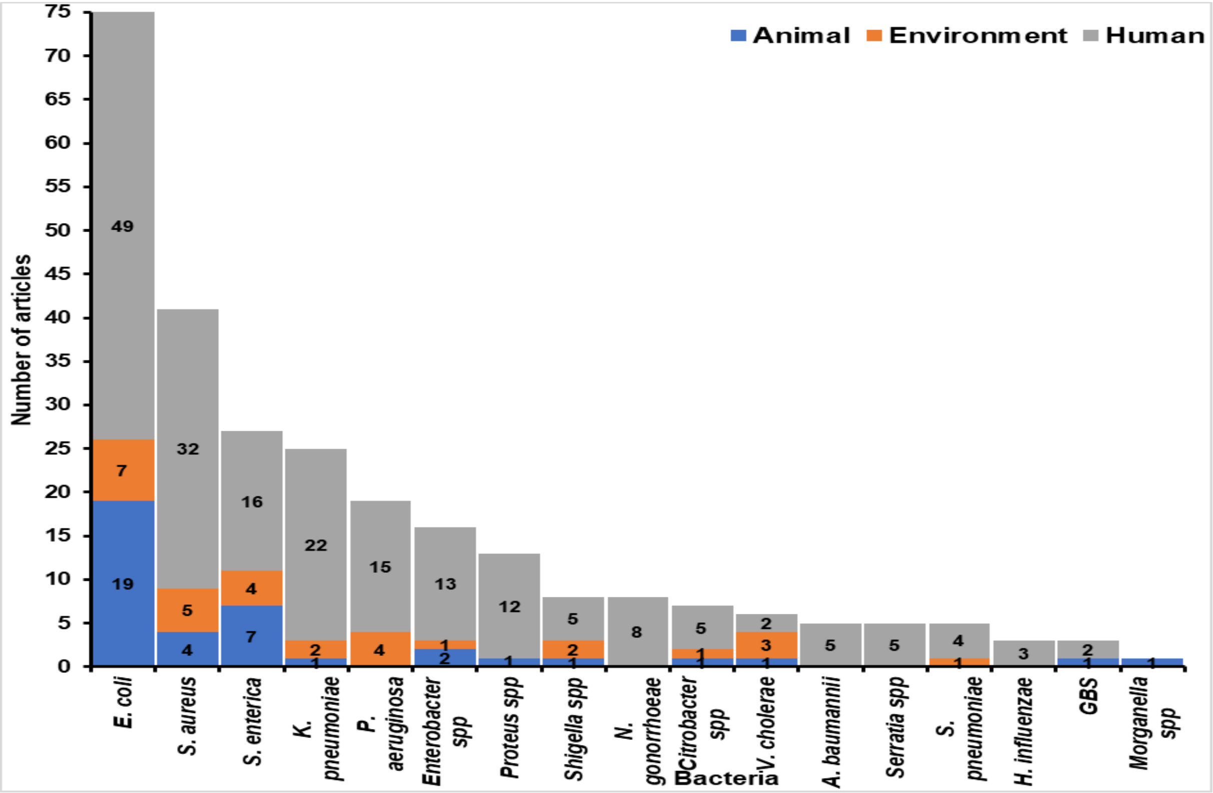


**Supplementary figure 3:** Frequency of studies reporting 17 priority bacteria in Cameroon.

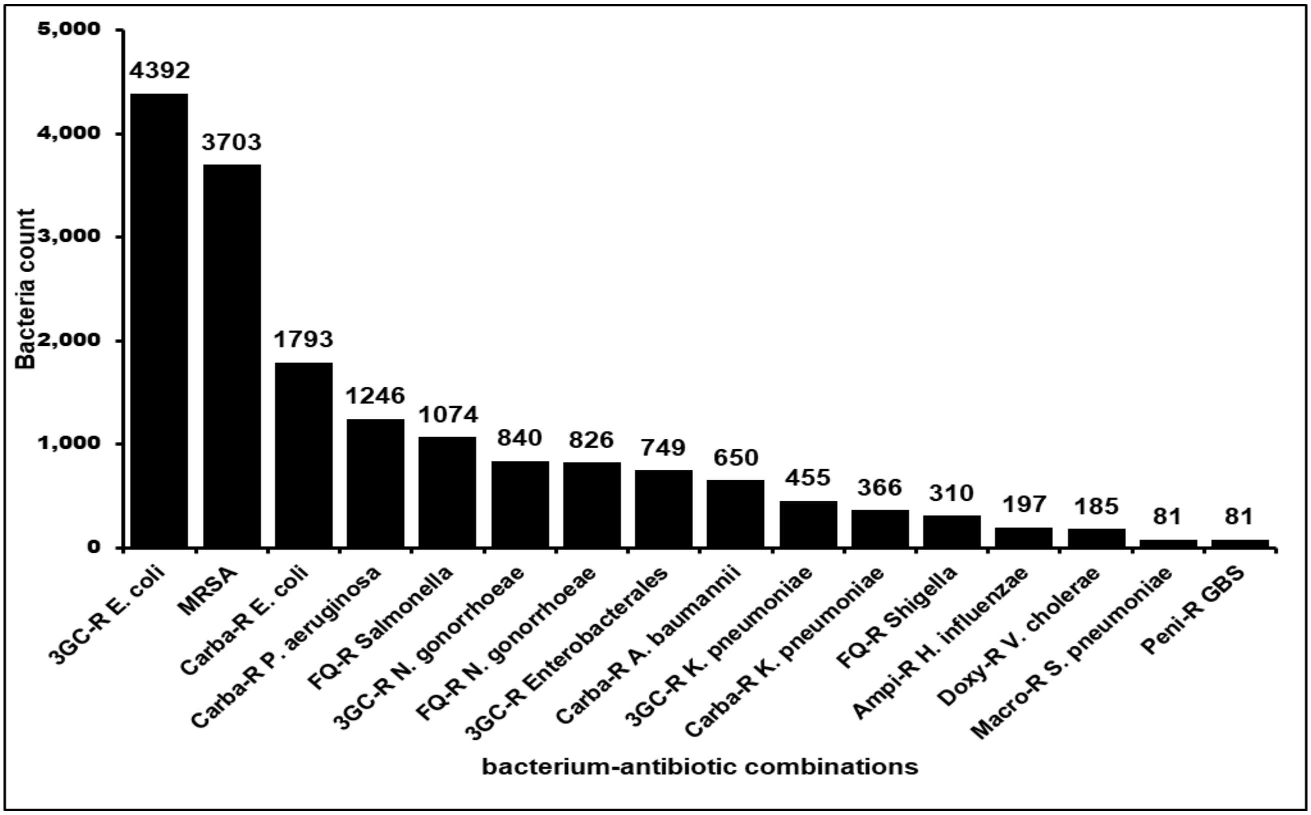


**Supplementary figure 4:** Frequency of WHO priority bacteria-antibiotic combinations reported. 3GC-R=Third-generation Cephalosporin Resistant, MRSA=Methicillin Resistant *S. aureus*, Carba-R=Carbapenem-Resistant, FQ-R=Fluoroquinolone-Resistant, Ampi-R=Ampicillin-Resistant, Doxy-R=Doxycycline-Resistant, Macro-R=Macrolide-Resistant, Peni-R=Penicillin-Resistant.

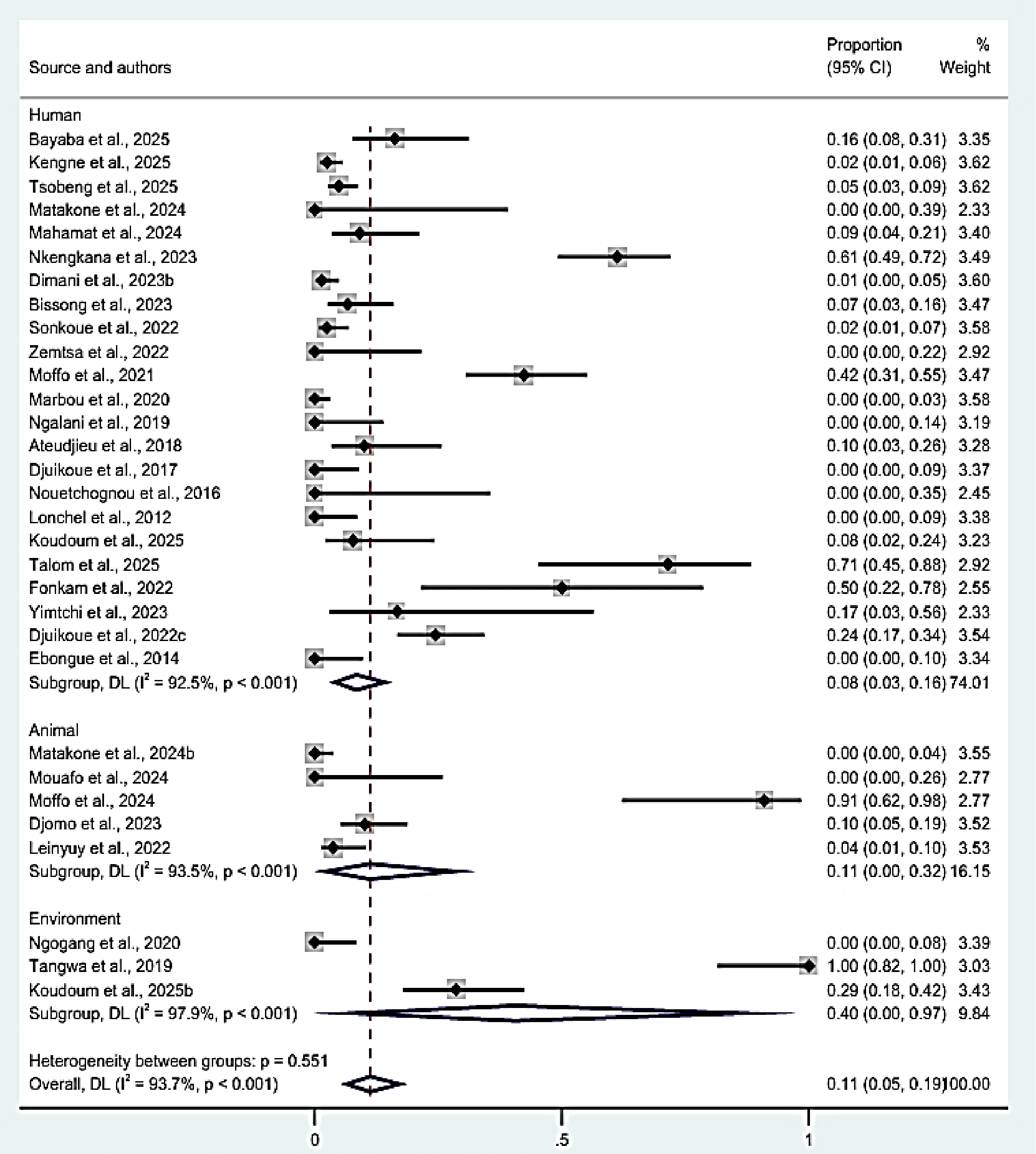


**Supplementary Figure 1:** Forest plot of pooled prevalence of carbapenem resistance in *E. coli* with subgroup meta-analysis by source. Note: weights and between subgroup analysis are from random effect models.

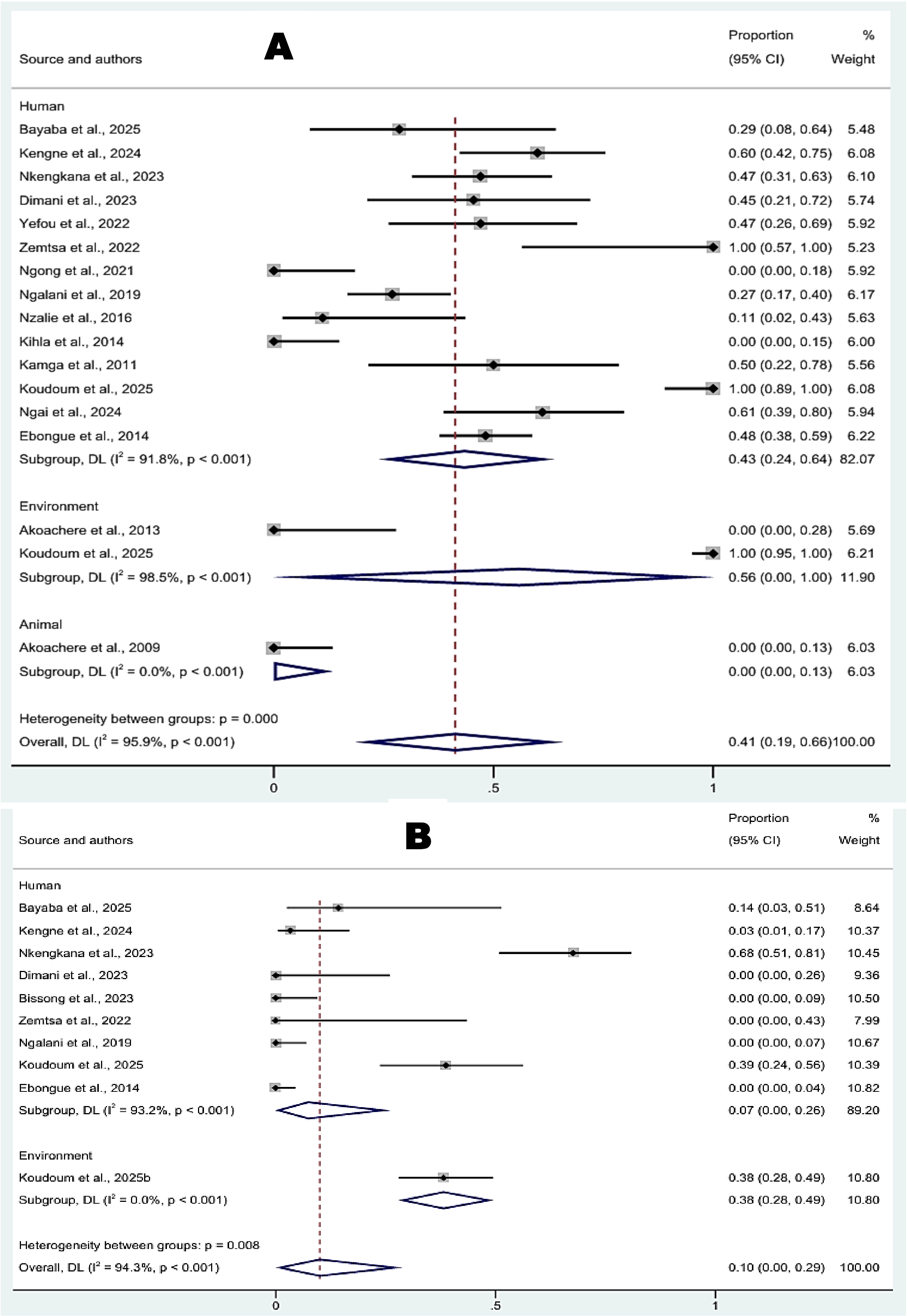


**Supplementary Figure 2:** Forest plot of pooled prevalence of third generation (A) and carbapenem (B) resistance in *K. pneumoniae* with subgroup meta-analysis by source. Note: weights and between subgroup analysis are from random effect models.

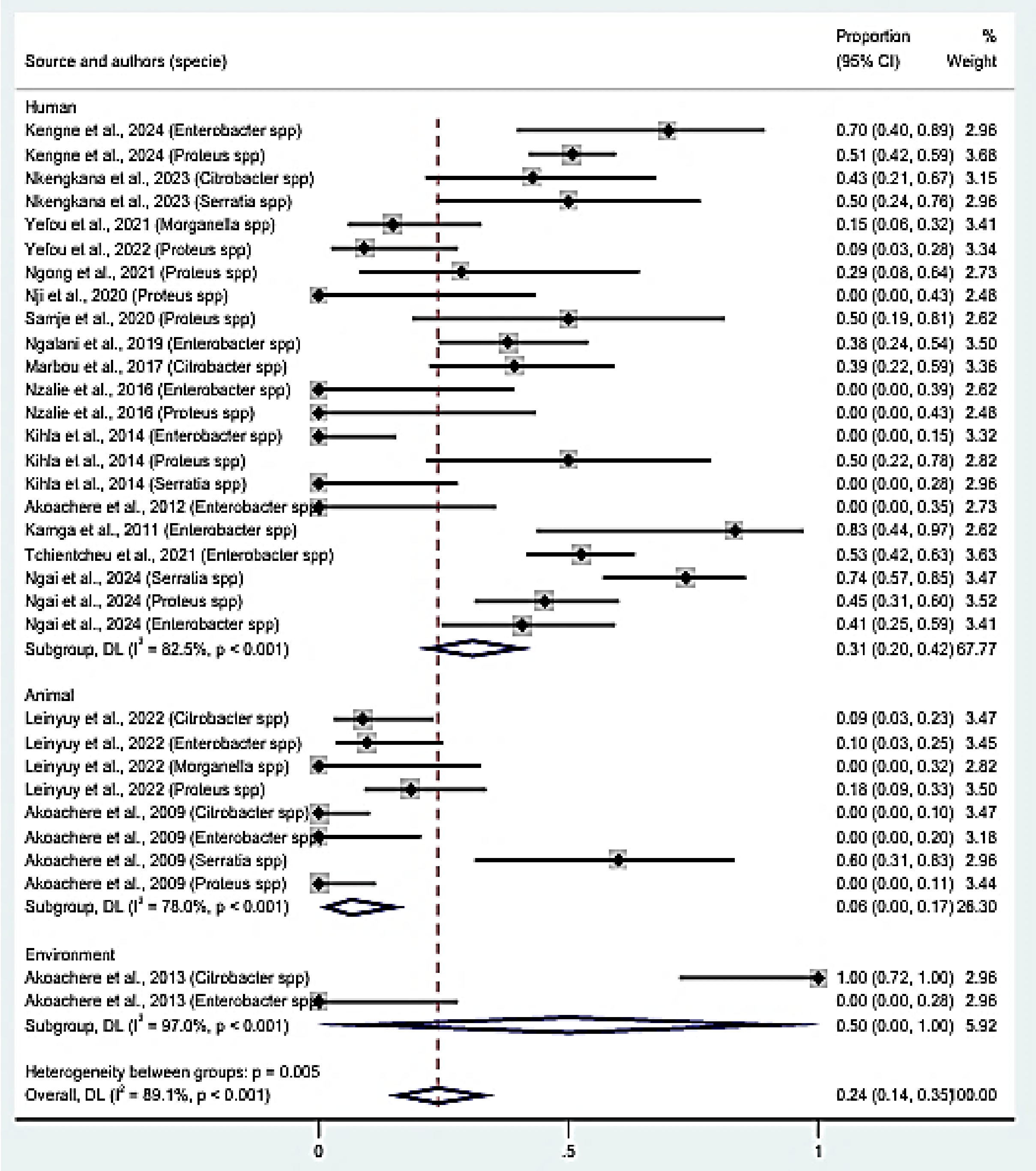


**Supplementary Figure 3:** Forest plot of pooled prevalence of third generation resistance in other *Enterobacterales* with subgroup meta-analysis by source. Note: weights and between subgroup analysis are from random effect models.

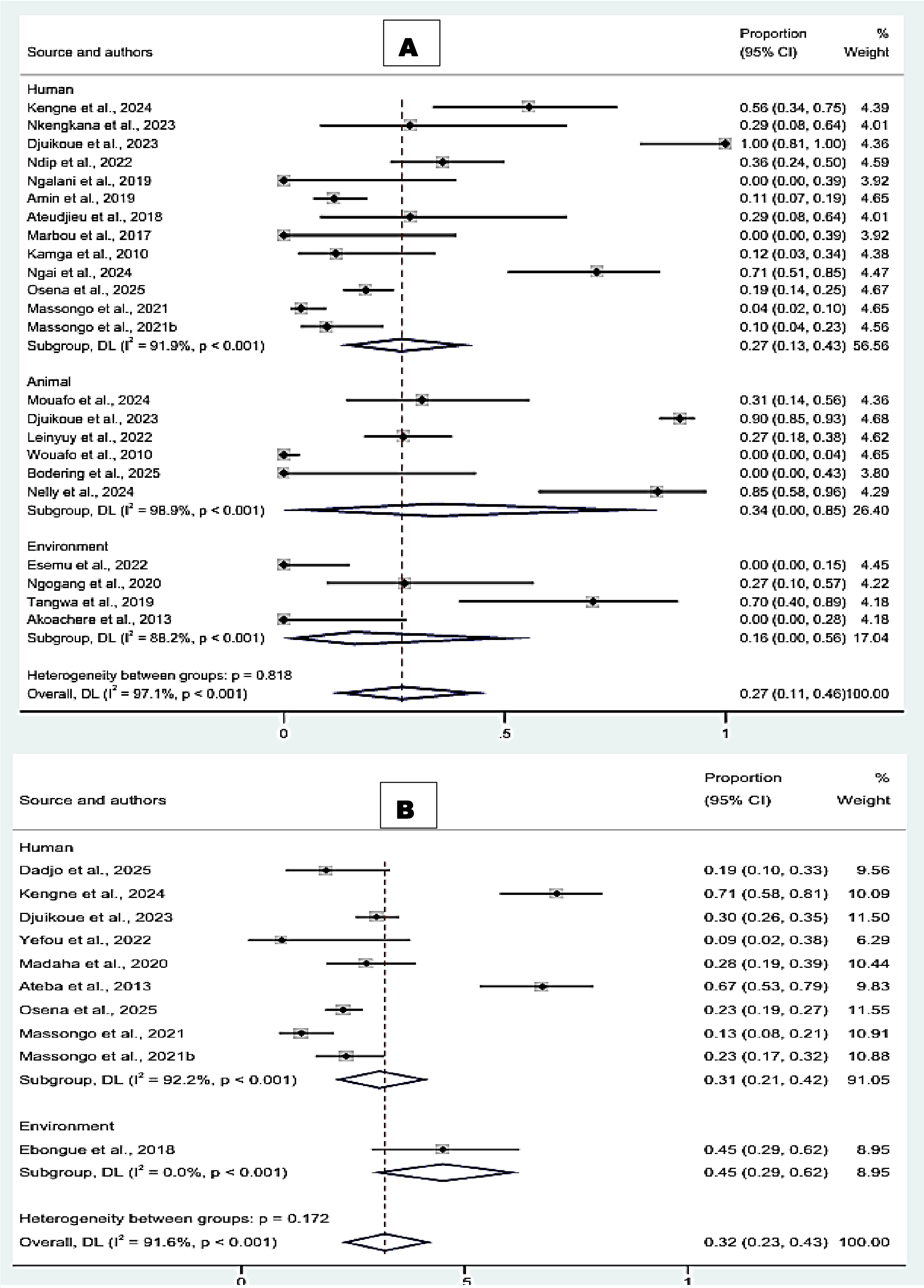


**Supplementary Figure 4:** Forest plot of pooled prevalence of fluoroquinolone resistance in *Salmonella* spp. (A) and Carbapenem resistance in *P. aeruginosa* (B) in with subgroup meta-analysis by source. Note: weights and between-subgroup analysis are from random effect

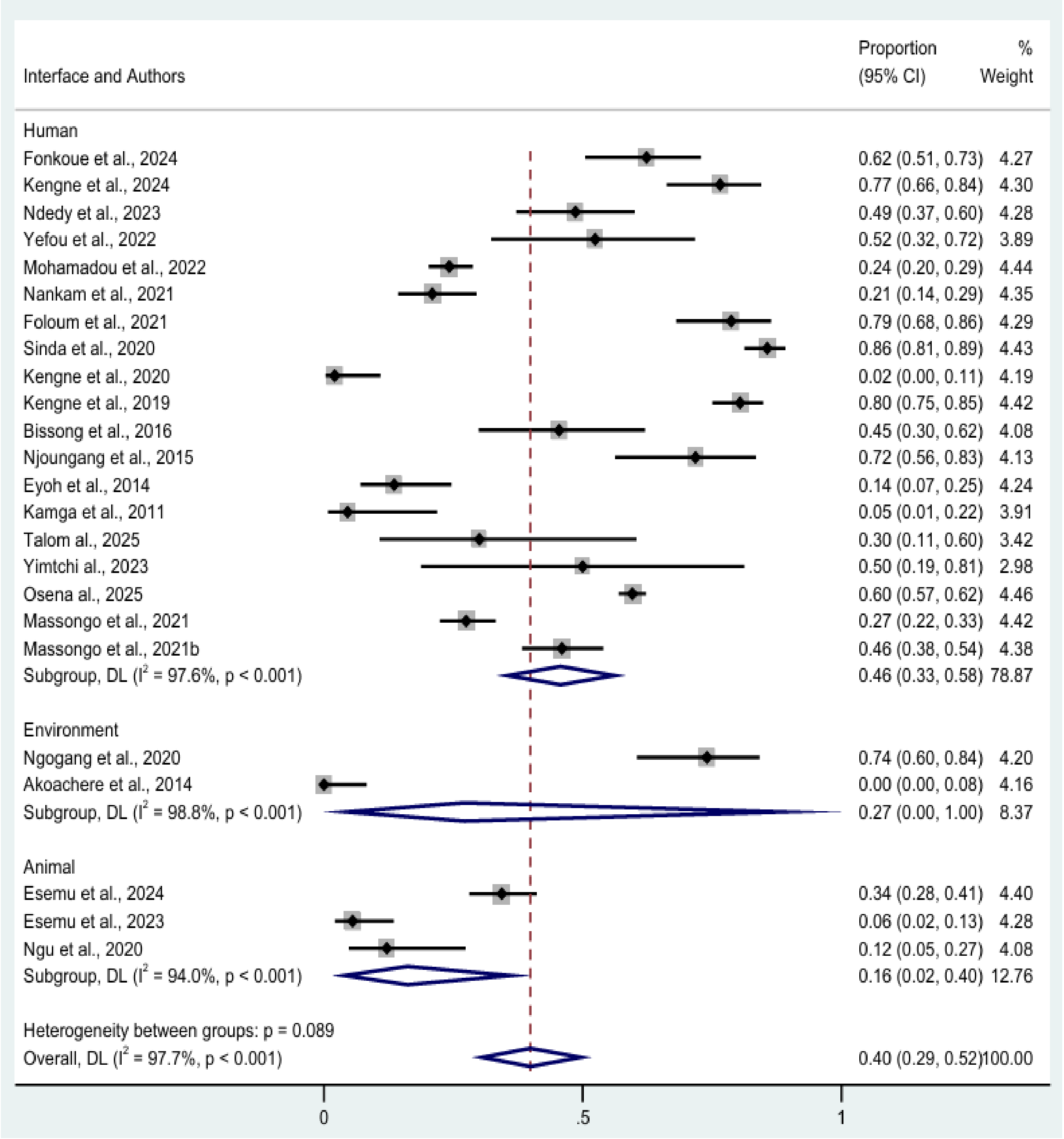


**Supplementary Figure 5:** Forest plot of pooled prevalence of methicillin resistance in *S. aureus* with subgroup meta-analysis by source. Note: weights and between subgroup analysis are from random effect models.

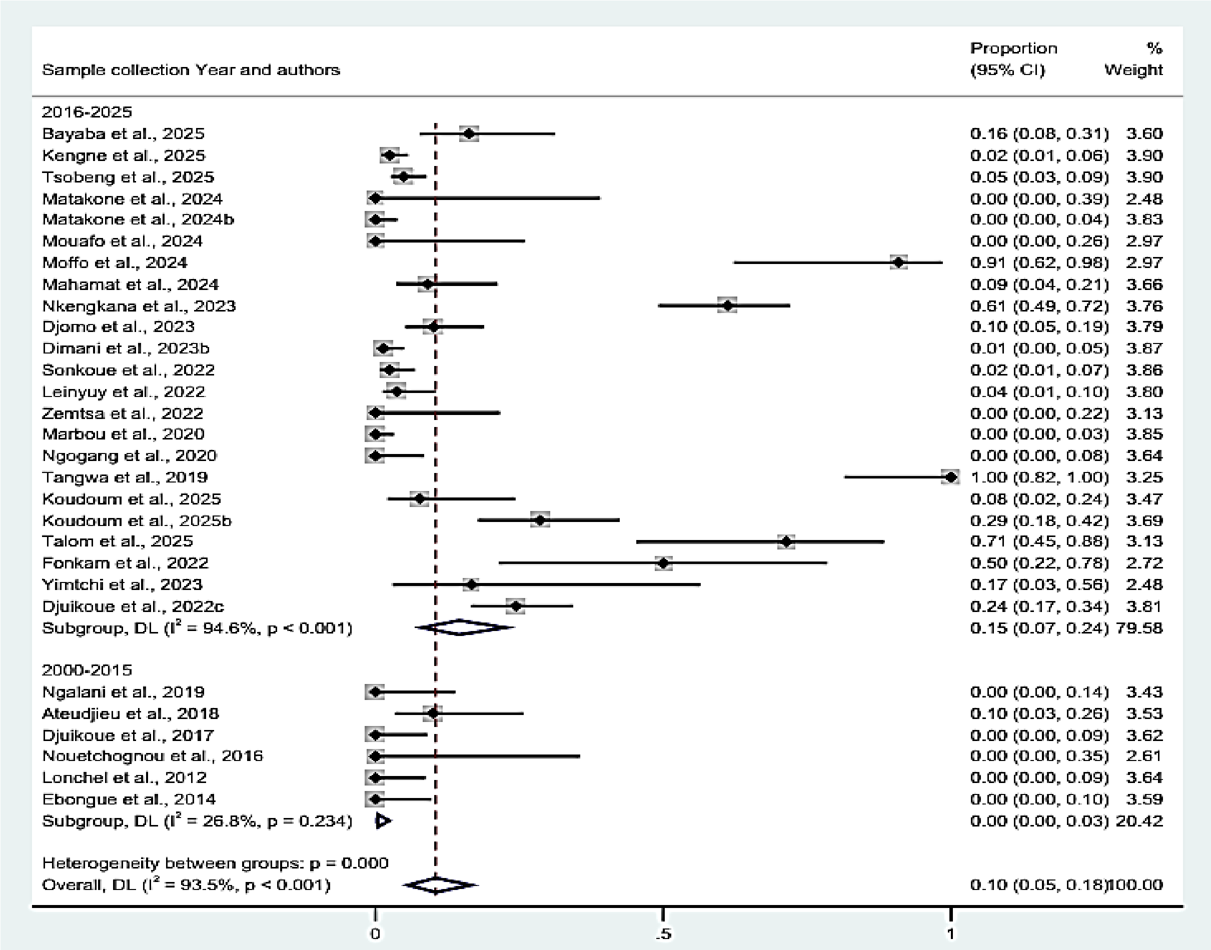


**Supplementary Figure 10:** Variation of pooled prevalence of carbapenem resistance in *E. coli* according to sampling year.
